# Magnitude of nonadherence to diet and exercise recommendations and associated factors among type 2 diabetes patients on treatment follow-up at Asella Referral and Teaching Hospital, Arsi, Ethiopia: A cross sectional study

**DOI:** 10.1101/2025.08.05.25333094

**Authors:** Nebyu Mekonen, Zewdu Hurisa, Ayalneh Demissie, Tesfa G/meskel, Enku Selamu

## Abstract

**Background:** Type 2 diabetes mellitus is a major public health problem worldwide. Nonadherence to lifestyle modifications in individuals with type 2 diabetes poses a great problem.

**Objective:** This study assessed magnitude and factors associated with nonadherence to diet and exercise.

**Methods:** An institution-based cross-sectional study was conducted on 302 type 2 diabetic patients at Asella Referral and Teaching Hospital of Arsi University. The data were collected via structured questionnaire. Multivariate logistic regression was used to determine the effects of independent variables on nonadherence to diet and exercise recommendations.

**Results:** Of the individuals in the study, 247(81.8%) patients did not follow the exercise recommendations, and 157(52%) patients did not follow the diet recommendations for type 2 diabetes. Duration since diagnosis > 5 years (AOR = 2.1, 95% CI [1.18–3.36]), doctors’ advice (AOR= 2.3, 95% CI [1.24–4.47]) and family history of DM (AOR= 4.3, 95% CI [2.55–7.29]) were independent determinants of nonadherence to dietary recommendations. Similarly, female sex (AOR= 3.6, 95% CI [1.60–8.60]), duration > 5 years since the diagnosis (AOR= 2.1, 95% CI [1.03–4.43]) and educational status attending only primary school (AOR= 3.33, 95% CI [1.33– 8.34]) and only reading and writing or illiteracy (AOR= 6.7, 95% CI [1.86–24.29]) were independently associated with nonadherence to the exercise recommendation of type 2 diabetes patients.

**Conclusions:** Overall, in our study, many patients were nonadherent to diet and exercise recommendations. Female patients and patients who attended primary school or less and > 5 years since diagnosis were more likely to be nonadherent to exercise. The determinants of diet nonadherence were a lack of doctors’ advice, > 5 years since diagnosis and a lack of family history.

## INTRODUCTION

Diabetes mellitus (DM) refers to a group of metabolic diseases characterized by hyperglycemia (high blood glucose) resulting from defect in insulin secretion, insulin action, or both. Chronic hyperglycemia in diabetes is associated with long-term damage, dysfunction and failure of various organs especially eyes, kidneys, nerves, heart, and blood vessels (1).Type 2 diabetes mellitus (T2DM) is the most common form of diabetes in adults, accounting for 90% of all diabetic adults (2).

Over the past three decades, the number of people with diabetes mellitus has more than doubled globally, making diabetes mellitus one of the most important public health challenges in all nations. Type 2 diabetes mellitus (T2DM) and prediabetes are increasingly common among children, adolescents and younger adults (3). According to a population-based study performed in 81 different countries, 439 million adults are expected to be diagnosed with diabetes by 2030. Between 2010 and 2030, there will be a 69% increase in the number of adults with diabetes in developing countries and a 20% increase in developed countries (4).

In SubSaharan Africa, the incidence of Noncommunicable diseases (NCDs) is predicted to exceed that of infectious diseases by the year 2030. The number of people with diabetes is projected to increase to 300 million by 2025 and 366 million by 2030 from 171 million in 2000 (5,6).

According to a systematic review and meta-analysis, the overall magnitude of T2DM in Ethiopia is approximately 5.95% for patients aged ≥ 40 years and 3.6% for patients aged less than 40 years (3).

Oxidative stress, as well as inflammation, plays a critical role in the pathogenesis and progression of diabetes and diabetes-associated morbidity. Emerging evidence suggests that exercise training activates the expression of cellular antioxidant systems but is also able to produce ROS, which are by no means detrimental. Instead, they are required for normal force production in skeletal muscle, for the development of training-induced adaptations in endurance performance, and for the induction of endogenous defense systems (7,8). Regular exercise is associated with lower levels of inflammatory markers and, simultaneously, with increases in anti-inflammatory substances. Therefore, regular and moderate exercise training can have antioxidant and anti-inflammatory systemic protective effects on type 2 diabetes patients (9).

In patients with established type 2 diabetes, physical activity improves glycemic control and reduces visceral adipose tissue and plasma triglyceride levels (10). Physical activity also has a beneficial effect on glycemic control by increasing tissue sensitivity to insulin (11). Healthy dietary habits, such as eating foods high in fiber and whole grain but low in fats, sugars, and carbohydrates, can help decrease the blood glucose level and subsequently reduce the amount of insulin needed (12).

American diabetic association (ADA) suggests that people with type 2 diabetes should perform at least 150 min per week of moderate to vigorous aerobic exercise for at least 3 days during the week, with no more than two consecutive days between bouts of aerobic activity(1).

Diets rich in whole grains, fruits, vegetables, legumes, and nuts; moderate in alcohol consumption; and low in refined grains, red or processed meats, and sugar-sweetened beverages have been shown to reduce the risk of diabetes and improve glycemic control and blood lipids in patients with diabetes. With an emphasis on overall diet quality, several dietary patterns, such as Mediterranean, low glycemic index, moderately low carbohydrate, and vegetarian diets, can be tailored to personal and cultural food preferences and appropriate caloric needs for weight control and diabetes prevention and management (13).

Lifestyle modifications among patients with T2DM are effective if healthcare practitioners understand patients’ reasons for adherence and nonadherence to diets and exercise recommendations (14). Certain studies indicate that adherence to a prescribed diet and regular exercise are important for both the prevention and control of patients with type 2 diabetes mellitus (15,16).

Adherence to prescribed lifestyle changes has also been shown to improve glucose levels, decrease blood pressure and correct lipid abnormalities, which are factors associated with the micro- and macrovascular complications of diabetes (17).

A research project at the Internal Medicine Clinic of Shebin El-Kom Teaching Hospital in Menoufia Governorate revealed that approximately 17% of participants engaged in physical activity with 50% of those who exercised reported doing so at least three times weekly, with sessions typically lasting 10–19 minutes or longer. However, the majority of the study subjects did not maintain a regular exercise routine. The mentioned obstacles to consistency were time constraints, social disapproval, unfavorable weather conditions, and exhaustion. With respect to adherence to healthy dietary habits, more than half of the studied subjects adhered to dietary recommendations. However, more than one-third of the studied subjects did not adhere to their diet, and the reasons for this were financial constraints, poor self-control, situations at home and the need for self-permissions (17).

In a systemic review performed in Africa, the most commonly explained barriers to poor adherence to lifestyle intervention, in order of importance, included a lack of knowledge and education, poverty and cost, population changes, and lack of access to healthcare (18).

A cross sectional study performed in four provinces of Kenya in the year 2010 revealed that 75% of the people who participated in the study had poor dietary practices, whereas 72% did not participate in regular exercise, and approximately 80% did not monitor their body weight (19).

In a study performed at Debre Tabor General Hospital, Northwest Ethiopia, a significant percentage (74.3%) of the study participants had poor adherence to dietary recommendations. The study reported that participants had a low consumption of fruits and vegetables and a low consumption of foods high in omega-3 fats, with means of 1.84 ± 1.96 and 0.1 ± 0.62 times a week, respectively. The listed reasons on the study for poor adherence to diet were lack of knowledge, lack of diet education, inability to afford the cost of a healthy diet and poor awareness of the benefits of dietary recommendations. The listed factors associated with nonadherence were, a low educational status, the presence of comorbidities, lack of previous dietary education and low monthly income (20).

In a study performed at Tikur Anbessa Specialized Hospital, Ethiopia 318 (75.9%) diabetes patients did not adhere to the recommended diet management, and 194 (46.3%) of the study subjects adhered to physical exercise, which means that they performed at least 30–60 minutes of moderate aerobic activity per day or ≥3 days per week. The majority of respondents, 104 (53.6%), were physically exercising for 30–45 minutes per session (21).

In a study performed at Police Hospital, Addis Ababa reported that Half (50.1%) of the study participants had poor adherence to common recommendations (diet, exercise, and medication). Specifically, most (53.7%) of the participants had good adherence to their diet, although it was lower than their adherence to exercise (87.5%) and medications (87.3%). The listed reasons associated with poor adherence to particular recommendations were a lack of knowledge (68.9) as a major reason for poor dietary adherence, whereas a lack of knowledge was the least likely reason for poor adherence to exercise (2.2%). In the study (37.8%) of the participants reported carelessness as their main cause of poor adherence to exercise (22).

A study performed in Illuababor, southwest Ethiopia, revealed that 38% of diabetic patients were nonadherent to physical activity recommendations. The odds of nonadherence to physical activity recommendations were independently associated with patient sex (AOR=2 (95% CI: 1.2, 3.4)), perceived severity of the illness (AOR=1.7 (95% CI: 1.1, 2.8)) and self-efficacy (AOR=2.6 (95% CI: 1.6, 4.4)) (23).

In a study performed in southwest Ethiopia, more than one-third (36.0%) and almost two-thirds (64.3%) of the patients did not adhere to diet and physical activity recommendations, respectively. Male sex, chewing khat, lack of social support and not receiving doctor recommendations were associated with nonadherence to dietary recommendations. Moreover, being female, being > 60 years of age, being illiterate, lacking social support and having disease duration > 5 years were associated with non-adherence to exercise recommendations (24).

In ARTH, we have many patients who visit diabetic clinics with T2DM who are non-adherent to lifestyle modifications, such as exercise and dietary recommendations, despite interventions such as advice and educational posters in the clinic, still have many patients with poor adherence. The problem could be patient related factors or the effectiveness and magnitude of the interventions performed which was not studied in ARTH.

Assessing adherence to diet and exercise recommendations in T2DM patients will help identify the magnitude and associated factors in local areas enabling targeted interventions such as exercise programs and dietary interventions and if modifiable factors such as a lack of advice from physicians and nursing staff are noted, we can intervene with teaching programs to aid in increasing overall adherence to lifestyle modifications for patients with T2DM.

The results of this study will help health care providers provide education and awareness on diet and exercise interventions targeting specific groups of patients in the management of DM. Thus, patients receive appropriate care that fits them. Therefore, this study was conducted to assess the magnitude of non-adherence to diet and exercise recommendations for T2DM and associated factors among patients with T2DM on follow up at ARTH diabetic clinic.

## MATERIALS AND METHODS

### Study setting and period

The study was conducted at Asella Referral and Teaching Hospital. The hospital is found in Asella, which is the seat of the East Arsi zone, Oromia. The town is located 175 km southeast of the capital Addis Ababa. The ARTH serves a catchment population of more than 3.5 million people from Oromia regional states in the country. The hospital provides inpatient and outpatient services. In addition, this ARTH medical college teaches under graduate and postgraduate medicine and other health science students.

The ARTH diabetic follow-up clinic has a total of 2,463 patients, 1,500 of whom have T2DM. The clinic has 3 nursing staff members. Medical residents and internists work in the clinic all week. The study was conducted from January 1 to January 30, 2025

### Study design

An institution based cross-sectional study was conducted to assess the magnitude and associated factors of nonadherence to diet and exercise modifications among T2DM patients on treatment follow up at the ARTH diabetic clinic.

### Sample size and sampling procedure

The minimum sample size was calculated using Cochran’s formula: n = Z^2^pq/d^2^ [12] at a prevalence (p) of 64.3% and 36% to nonadherence to diet and exercise recommendations based on a study performed in Jimma Medical Center, respectively (24). By taking the higher prevalence to get the maximum sample size: p = 0.643 and q = 1 – 0.643=0.357 and d, the tolerated margin of error =5% at the 95% confidence level (25).

Thus, (1.96)^2^ (0.643) (0.357)/0.05×0.05 = 352.73724864 ∼353 patients. Since the target population is less than 10,000, we used the correction formula nf = No/ 1 + No/N, where nf is the corrected sample size and N is an estimate of the population size during the study period which is 1500. The corrected sample size was nf = 353/1 + (353)/1500 = 286 and by adding a 10% contingency for non-responses, the final sample size was 315 patients.

Patients with type 2 diabetes who attended the diabetic clinic at ARTH for regular follow-up visits were included in the study but patients who are acutely ill and can’t respond to questions; patients with impaired memory or who can’t respond to the questions and patients with disability were excluded from the study.

Systemic random sample was used to select the required number of T2DM patients, taking every fourth patient (1,500/315∼4.7 based on a visiting sequence) using the diabetic clinic follow-up record as a sampling frame.

### Data collection tools

The data were collected face to face using interviewer administered questionnaire that developed after reviewing different literature on related topic (23,24) by two trained nurses at the diabetic clinic and supervised by master graduate public health professionals. The information collected included sociodemographics, behavioral, environmental and clinical information. The field work was conducted from January to February 2025. T2DM patients were interviewed at clinic by trained data collectors who were fluent in Afan Oromo following sensitivity analysis. Each interview was carried out face-to-face and lasted approximately 20 minutes. Patients were be classified as nonadherent if they didn’t fulfill the requirement criteria and adherent if they fulfilled the requirement criteria.

### Data quality control

The questionnaire was first developed in English and translated to Afan Oromo (the regional state language) by language expert and re-translated back to English to check its consistency. Before conducting the actual study, the questionnaire was pre-tested for validity and reliability on 5% of sample size in Adam Hospital Medical College which is located 75 kilometers away from the study hospital and modification of the data collection tool was made on the logical sequence of the questionnaire, its grammar and how to conduct interview. Training was given for data collectors and supervisors for one day by principal investigator about the objective of the study and data collection methods. During the actual data collection, close supervision was made by the supervisors and principal investigator on a daily basis to check consistency and completeness of questionnaire.

### Measurements

The outcome variables were nonadherence to diet and exercise recommendations. Nonadherent to exercise: If a patient does not engage in moderate intensity exercise for at least 150 minutes per week and at least three times per week. It can be mixed with strength exercise or 75 minutes of vigorous intensity exercise (1). Nonadherent to diet: If a patient uses simple sugar and sweetened beverages and simple starch (pasta, rice, white bread, cookies or cake), does not eat fruits or vegetables at least three times per week(26).

The determinants considered in the study included demographic and clinical characteristics, and a number of behavioral variables. . We examined the explanatory value of a number of socio-demographic variables included age, sex, marital status, children status, educational status, residence, monthly and income. Then we examined respondents’ clinical variables: duration of the disease, family history of diabetes, doctors’ advice and comorbidity. Lastly the study assessed behavioral variables: BMI, social support, cigarette smoking, Khat chewing and alcohol consumption. Moderate alcohol intake: One drink per day for female and 2 drink per day for male (27). One drink equals 12 grams of ethanol) which is equivalent to one bottle of bear, a glass of wine, (27) 40 ml areke, 350 ml tella and 200 ml tej (28).

### Data analysis

The data were analyzed using stata version 25. Descriptive statistics such as percentage and frequency were computed, and the mean with standard deviation was used to summarize the continuous variables accordingly. Both bivariate and multivariable logistic regressions were performed to identify significant factors associated with adherence. Those variables with p-value < 0.25 in bivariate analysis were considered for multivariable logistic regression analysis to control the effect of confounding variables. Multicollinearity was checked among independent variables using variance inflation factors, and then the highest VIF between the independent variables was found to be 1.299, which was below cut-off points. The model fitness was checked by using the Homer-Lemeshow goodness of fit test and found to be p-value = 0.551.The strength of associations was expressed using adjusted odds ratios (AORs) with 95% confidence intervals, and the significance of associations was declared at a p-value of less than 0.05. Finally, the results are presented using charts and tables.

### Ethical considerations

Ethical approval was obtained from Arsi University Ethical Review Committee (A/CHS/RC/138/2024) and permission letter was obtained from ARTH before field activities started. After a detailed explanation of the purpose, benefits, confidentiality of the information, and voluntary nature of participation in the study we obtained written informed consent from all participated T2DM patients prior to their participation. Name and other personal identifiers were not recorded to maintain confidentiality.

## RESULTS

### Sociodemographic characteristics of the respondents

Interviews with 302 of the 315 patients who were required were completed successfully (response rate of 96%). Among the participants, 171 (56.6%) were male. A total of 161 (53.3%) of them were aged above 50 years. Most are married 204 (67.5%) and the majority (91.7%) had children. A total of 55.3% resided in rural areas. The educational background of the participants revealed that 109 (36.1%) had attained only primary school education, 98(32.4%) had completed secondary school or above and 31.4% only read and write or illiterate, as shown in table 1.

**Table 1:** Demographic characteristic of the participants in ARTH, 2025. (n=302)

| Variable | Category | Frequency | Percentage |
| --- | --- | --- | --- |
| Age | 30-50 | 141 | 46.7 |
|  | Above 50 | 161 | 53.3 |
| Sex | Female | 131 | 43.4 |
|  | Male | 171 | 56.6 |
| Marital | Single | 27 | 8.9 |
|  | Married | 204 | 67.5 |
|  | Divorced | 33 | 10.9 |
|  | Widowed | 38 | 12.6 |
| Children | No | 25 | 8.3 |
|  | Yes | 277 | 91.7 |
| Residence | Rural | 167 | 55.3 |
|  | Urban | 135 | 44.7 |
| Education | Secondary school and above | 98 | 32.4 |
|  | Primary school | 109 | 36.1 |
|  | Illiterate and Only read and write | 98 | 31.4 |
| Occupation | Daily laborer | 10 | 3.3 |
|  | Employed | 48 | 15.9 |
|  | Farmer | 112 | 37.1 |
|  | Housewife | 73 | 24.2 |
|  | Other | 5 | 1.7 |
|  | Private business | 46 | 15.2 |
|  | Unemployed(not working) | 8 | 2.6 |
| Income in ETB* | < 5000 | 113 | 37.4 |
|  | 5000 – 10000 | 136 | 45 |
|  | > 10000 | 49 | 16.2 |

**ETB\* Monthly income in Birr**

### Clinical and Behavioral characteristics

Among the 302 participants, 54.6% had T2DM for more than 5 years. A total of 46% of the participants reported a family history of diabetes. A total of 52.3% of the participants utilized only oral hypoglycemic agents and 25.2% used only insulin, and the remaining 22.5% employed both. Twenty-one percent of the participants did not receive advice from health professionals regarding lifestyle modifications for T2DM. A total of 92.4% of the participants had social support. Only 14 (4.6%) and 12 (4%) patients chewed khat and smoked cigarettes, respectively, whereas 226 (74.8%) patients did not drink alcohol (Table 2).

**Table 2:** Clinical and behavioral characteristics of participants in ARTH, 2025. (n=302)

| Variable | Category | Frequency | Percentage |
| --- | --- | --- | --- |
| BMI | <18.5 | 26 | 8.6 |
|  | 24.9 - 18.5 | 226 | 74.8 |
|  | 29.9 - 24.9 | 50 | 16.6 |
| Duration | > 5 years | 165 | 54.6 |
|  | ≤ 5years | 137 | 45.4 |
| Comorbidity | No | 123 | 40.7 |
|  | Yes | 179 | 59.3 |
| Family history | No | 163 | 54 |
|  | Yes | 139 | 46 |
| Medication | Oral hypoglycemic agent | 158 | 52.3 |
|  | Insulin | 76 | 25.2 |
|  | Both oral hypoglycemic agent and insulin | 68 | 22.5 |
| Advice | No | 65 | 21.5 |
|  | Yes | 237 | 78.5 |
| Support | No | 23 | 7.6 |
|  | Yes | 279 | 92.4 |
| Alcohol | Above moderation | 23 | 7.6 |
|  | In moderation | 53 | 17.5 |
|  | No | 226 | 74.8 |
| Smoking | No | 290 | 96 |
|  | Yes | 12 | 4 |
| Khat | No | 288 | 95.4 |
|  | Yes | 14 | 4.6 |

Among the participants, 179 (59.3%) had comorbidities, with hypertension being the predominant condition in 155 (86.5%) and 67 patients (37.4%) having cardiac illness; 9 (5%) patients had asthma, one patient had stroke, and 17 (9%) patients had other comorbidities (chronic obstructive lung disease, chronic lower back pain).

### Magnitude of nonadherence to diet and exercise recommendations

Among the participants 247 of them (81.8%) were not adherent to exercise recommendations, and 157 (52%) of the participants were non-adherent to dietary recommendation of T2DM.

### Reasons for nonadherence to the Recommended Lifestyle Modifications

The most common reason for nonadherence to dietary recommendationd was cost of the recommended diet (47 patients). The most common reason for non-adherence to exercise was lack of interest (77 patients) followed by lack of knowledge (see figure 1 & 2).

**Figure 1:**
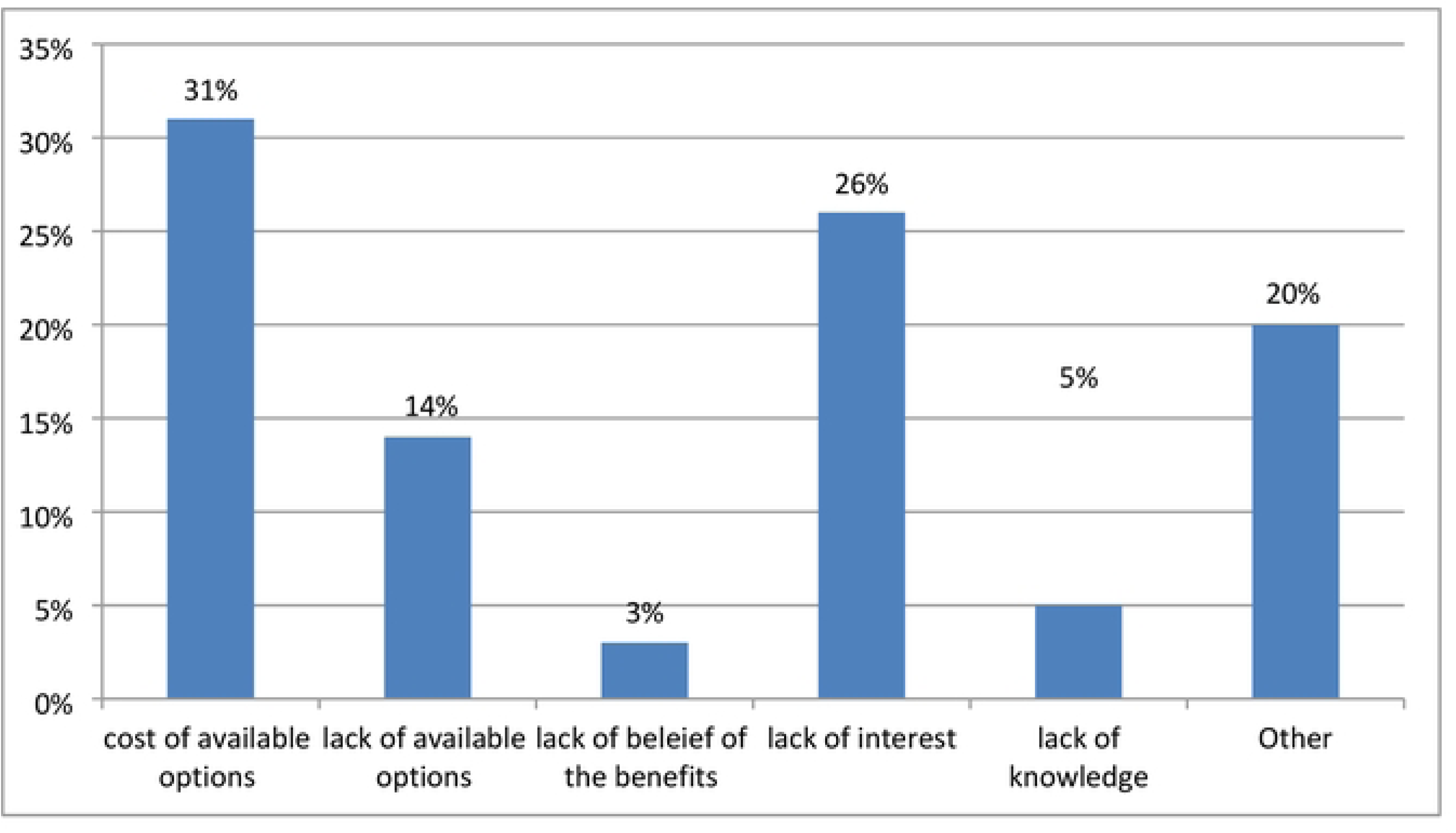
Reasons for nonadherence to dietary recommendations in ARTH, 2025. (n=152)

**Figure 2:**
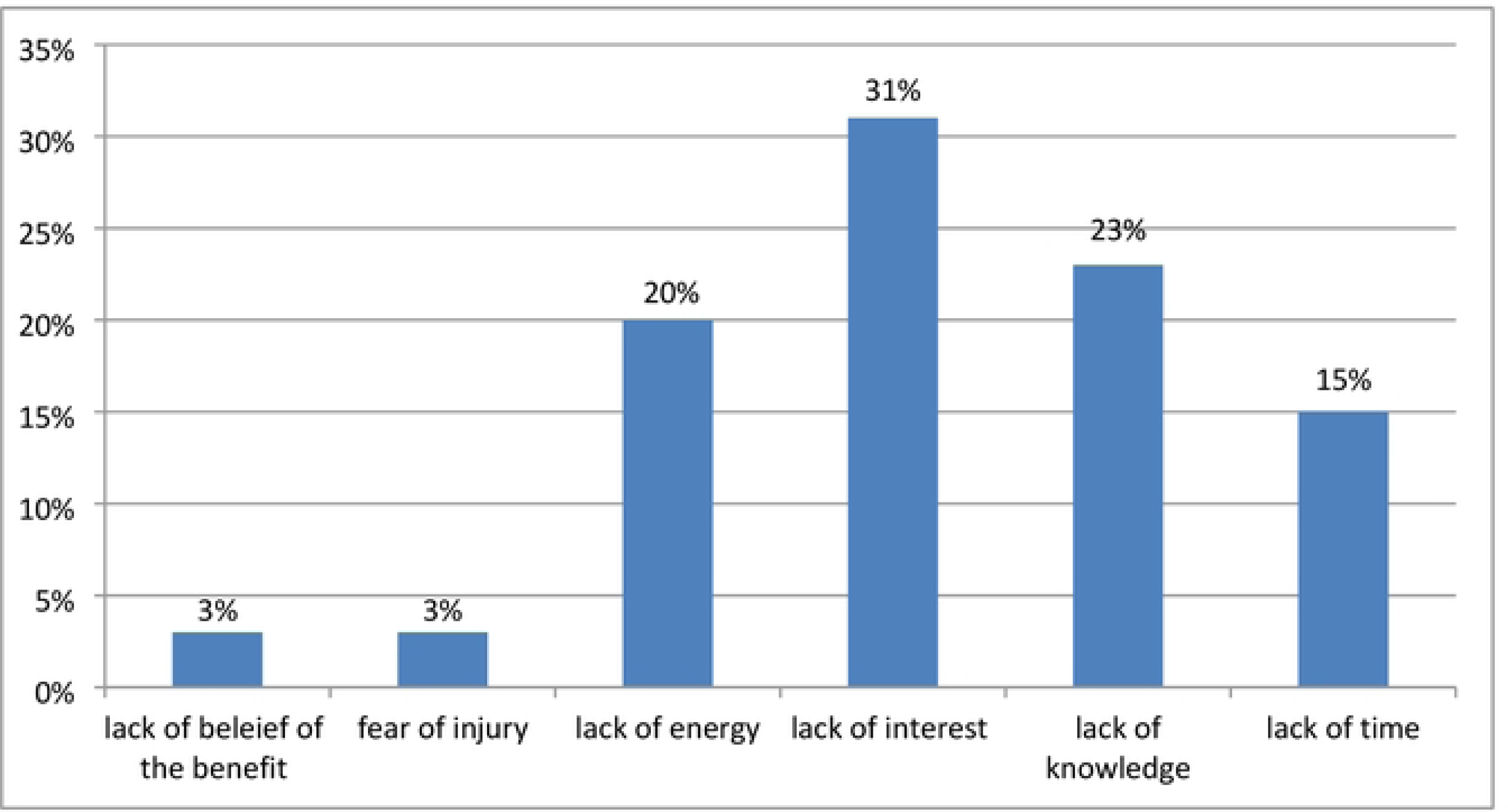
Reasons for nonadherence to exercise recommendation in ARTH, 2025. (n=247)

### Determinants of nonadherence to the Recommended Lifestyle Modifications

#### Determinants of nonadherence to exercise recommendations

Binary logistic regression was performed to identify associations between the baseline characteristics of the patients and nonadherence to exercise recommendations. Sex, residential marital status, educational status, having children, and duration since diagnosis had p values < 0.25. After that, multivariate regression was performed to identify independent determinants of nonadherence to exercise recommendations, after which female sex (AOR= 3.6, 95% CI [1.60-- 8.60]), duration ≥ 5 years since diagnosis (AOR= 2.1, 95% CI [1.03--4.43]) and educational status attending only primary school (AOR= 3.33, 95% CI [1.33--8.34]), only reading and writing and being illiterate (AOR= 6.7, 95% CI [1.86--24.29]) were independently associated with nonadherence to exercise recommendations for T2DM patients.

**Table 3:**
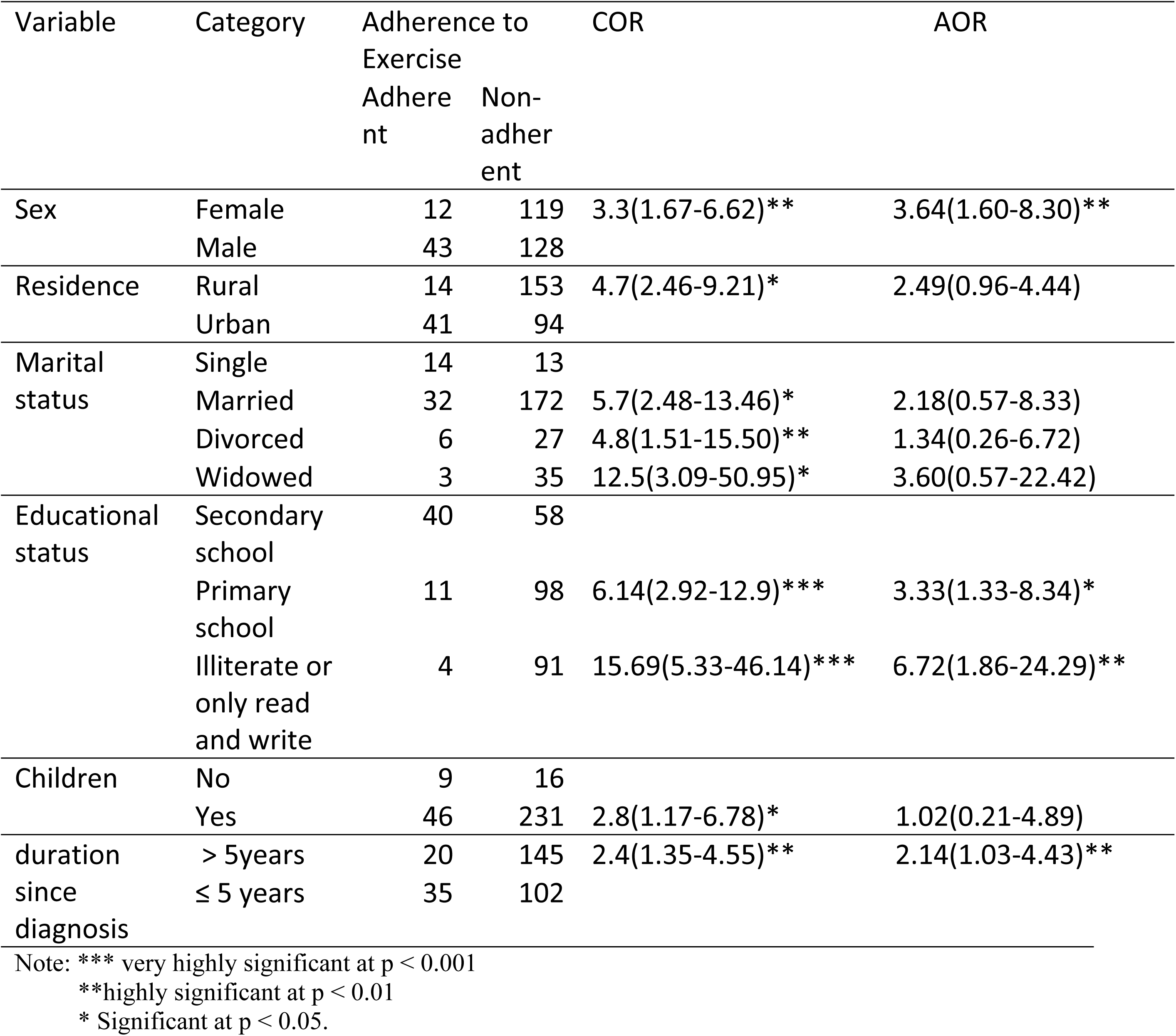
Bivariable and multivariable analysis of determinants significantly associated with nonadherence to exercise in T2DM in ARTH, 2025. (n=302)

### Determinants of non-adherence to dietary recommendation

Binary logistic regression was performed to determine the associations between baseline characteristics and nonadherence to dietary recommendations, from which age, alcohol consumption, cigarette smoking history, duration since diagnosis, advice from healthcare professionals, and social support had p values < 0.25.

Furthermore, multivariate logistic regression was performed to identify independent determinants of nonadherence to the dietary recommendation of T2DM, and the results revealed that duration since diagnosis more than 5 years (AOR= 2.1, 95% CI [1.18–3.36]), lack of advice from health professionals (AOR= 2.3, 95% CI [1.24–4.47]) and having no family history of DM (AOR=4.3, 95% CI [2.55–7.29]) were independent determinants of nonadherence to the dietary recommendations.

**Table 4:** Bivariable and multivariable analysis of determinants significantly associated with nonadherence to diet in T2DM in ATRH, 2025. (n=302)

| Variable | Category | Non-adherence to diet |  | COR(95%CI) | AOR(95%CI) |
| --- | --- | --- | --- | --- | --- |
|  |  | Adherent | Non-adherent |  |  |
| Age(1) | 30-50 | 86 | 55 |  |  |
|  | Above 50 | 71 | 90 | 1.98(1.25- 3.13)** | 1.53(0.85-2.77) |
| Alcohol | No | 129 | 97 |  |  |
|  | In moderation | 20 | 33 | 2.19(1.18-4.05)* | 2.19(1.07-4.48) |
|  | Above moderation | 8 | 15 | 2.49(1.01-6.11)* | 1.74(0.63-4.77) |
| Cigarette | No | 155 | 135 |  |  |
|  | Yes | 2 | 10 | 5.74(1.23-26.66)* | 2.96(0.53-16.47) |
| Advice | Yes | 132 | 105 |  |  |
|  | No | 25 | 40 | 2.01(1.147-3.52)* | 2.35(1.24-4.47)** |
| Family history | No | 58 | 105 | 4.48(2.75-7.29)*** | 4.30(2.55-7.29)** |
|  | Yes | 99 | 40 |  |  |
| Duration | >5 years | 70 | 95 | 2.36(1.48-3.76)*** | 2.14(1.18-3.36)* |
|  | ≤ 5years | 87 | 50 |  |  |

Note: \*\*\* very highly significant at $p < 0.001$
\*\*highly significant at $p < 0.01$
\* Significant at $p < 0.05$ .

## DISCUSSIONS

### Magnitude of nonadherence to diet and exercise recommendations Magnitude of nonadherence to dietary recommendations

In this study approximately half (52%) of the participants were nonadherent to dietary recommendations which is higher than study done in Egypt (33%) (14) possibly due to cultural and geographic differences and also higher than that reported in Jimma (36%) where the participants were considered adherent if the patient was adherent to avoid simple starch three times per week (24) which is different from our study. Similarly, a study at Addis Ababa police hospital reported a lower percentage of non-adherence (46.3%) which measured low sugar subjectively (22). In contrast, this percentage was lower than what was reported in the Tikur Anbessa Specialized Hospital study done in Addis Ababa (75.9%) (21). Different cultural eating habits may be the cause of this.

#### Magnitude of nonadherence to exercise recommendations

In this study, a Significant number of the participants (81.8%) were not adherent to exercise recommendation which is almost similar to finding from a study in Egypt (83%) (14) and higher than a study in Addis Ababa (53.7%) (21) which labeled patients adherent if they performed physical activity only three times a week for thirty minutes session in contrast to this study which used the current ADA guideline (1). Jimma (64.3%) (24), which also took lower frequency even once per week as adherent. This number is also significantly higher than that reported in other studies at Addis Ababa police hospital (22.5%), where more than half of the participants were military (22) who could be adapted to performing exercise.

#### Determinants of nonadherence to diet and exercise recommendations Determinants of nonadherence to diet

In this study, one of the independent determinants of nonadherence to diet was a lack of advice about lifestyle modification, with similar results to those reported in other studies (14,20,24) which could be due to a lack of knowledge of some of the recommendations. The other determinant was having no family history of DM which was not analyzed in other studies in respect to adherence to both diet and exercise (21,24) but this finding could be explained by the fact that having a family history could increase the familiarity with the dietary habits of diabetic patients. Furthermore, patients with a duration of DM of more than 5 years since the time of diagnosis were more likely to be nonadherent than those with a diagnosis of ≤ 5 years, which is a similar trend to that reported in a study in Northwest Ethiopia (23) and could be explained by the fact that a longer duration of the disease could increase the likelihood of being fed up with following dietary recommendation.

In contrary to other studies (24) adherence to diet was not significantly associated with the number of patients who chewed in our study, which may have affected the analysis (13 patients), and there was no significant association between monthly income and the level of adherence, which may be due to the failure to report the exact amount of income due to perceived social norms.

#### Determinants of nonadherence to exercise

In this study, independent determinants of nonadherence to exercise recommendations for T2DM patients were that female patients were more likely to be nonadherent to recommendations of exercise, which is in line with the findings of a study in Jimma, which showed that females were twice as likely to be nonadherent (24) and Southwest Ethiopia (23) could be attributed to a culture of women staying at home with responsibility of raising children. Not attending secondary school or above was the other predictor that was also evident in a study done in Jimma(24) which can be related to a lack of knowledge of the recommended duration and benefit of exercise and individuals with a diagnosis of diabetes for more than 5 years were also more likely to be nonadherent to exercise than those with more recent diagnosis (≤ 5 years), which could be because patients with longer durations of illness might stop performing physical activity when they are accustomed to living with diabetes and being fed up with exercise.

There were no statistical difference in the state of adherence to exercise recommendations between those who received advice from health professionals and those who did not which could be due to a lack of appropriate advice on the exact time and benefit of the exercise recommendations.

## CONCLUSIONS AND RECOMMENDATIONS

### Conclusions

This study revealed that many of the patients are not adherent to diet and exercise recommendations especially exercise. We found that female sex, education level below secondary school and duration since diagnosis more than 5 years were independently associated with nonadherence to exercise. Regarding diet having no family history of DM, a lack of advice by health care professional and a duration more than 5 years since diagnosis of disease were the factors that independently predicted nonadherence,

### Recommendations

Health care professionals should give emphasis for these groups of patients mentioned above when managing and advising patients on lifestyle modification. Furthermore, as significant numbers of patients give reason of lack of knowledge as a cause of their nonadherence to exercise therefore, Physicians and nurses should provide specific and accurate guide on the recommended 150 minutes per week exercise recommendation during patients’ follow up visits.

### Limitations

This research is not without its limitations. As we are only assessing patients’ responses to the question some patients may not report the true answer to avoid confrontation in addition to recall bias as well as social desirability bias. Despite our best efforts to account for a wide range of potential confounders, residual confounding might still exist. While the study provides valuable insights for the Asella region, it is possible that the findings may not be applicable to other settings with different medical practices, healthcare systems, or demographics.

## Data Availability

The data is found in the text under supporting information

## Supporting information

**S1 file. Diabetic diet and exercise questionnaire**

(DOCX)

**S2 file. Raw data; cross sectional**

(XLSX)

## Acknowledgements

The authors would like to express their sincere appreciation to the data collectors and study participants

## Authors’ contributions

NM developed analyzed, interpreted and wrote the first draft. ZH and AD provided valuable comments on the writing of the manuscript. All the authors read and approved the final manuscript.

